# Genotype–Phenotype Correlation and Mutational Burden in Colombian Patients with Congenital Adrenal Hyperplasia

**DOI:** 10.1101/2025.09.15.25335820

**Authors:** Ana M. Perilla-Espinal, Valentina Zapata-López, Sofía Villada-Montoya, Carolina Jaramillo-Arango, Jennyfer Monroy-Espejo, Carolina Baquero Montoya, Cruz E. Zabala-Granda, Carolina Prieto-Saldarriaga, Gustavo A. Giraldo Ospina, Clara M. Arango-Toro, Carlos E. Builes-Montaño

## Abstract

**Background:** Congenital adrenal hyperplasia (CAH) due to 21-hydroxylase deficiency (21OHD) is characterized by a broad clinical spectrum, ranging from salt-wasting to nonclassical forms. Genotype–phenotype correlations based on predicted residual enzymatic activity have been widely studied, but data from Latin American populations remain scarce. Additionally, the influence of mutational burden on phenotype prediction has not been fully explored.

**Objective:** To evaluate the genotype–phenotype correlation and the impact of mutational burden on predictive accuracy in a Colombian cohort of patients with CAH.

**Methods:** We conducted a cross-sectional study of patients with confirmed CAH enrolled in a specialized rare disease program. Genotypic classification was based on predicted residual enzymatic activity (Null, A, B, C), and clinical phenotype was categorized as salt-wasting (SW), simple virilizing (SV), or nonclassical (NC). Genotype–phenotype concordance was defined as exact category agreement. Mutational burden was defined as the total number of pathogenic variants, dichotomized as low (≤2 mutations) or high (>2). Penalized logistic regression (Firth method) was used to evaluate associations between mutational burden, sex, and concordance.

**Results:** Among 48 patients with available genetic data, genotype–phenotype concordance was highest in severe genotypes: 100% in Null and 85.7% in Group A. In contrast, concordance declined in Group B (33.3%) and Group C (44.4%). Individuals with high mutational burden had significantly lower odds of concordance (OR = 0.18; 95% CI: 0.03–0.94). No significant interaction between sex and mutational burden was observed. More than one-third of Group C patients exhibited more severe phenotypes than predicted.

**Conclusions:** Our findings support established genotype–phenotype correlations in CAH, particularly for severe genotypes. However, increased mutational burden was associated with reduced predictive accuracy, suggesting the need to consider total mutation load in clinical assessment and genetic counseling.

## Introduction

Congenital adrenal hyperplasia (CAH) encompasses a group of autosomal recessive disorders caused by enzymatic deficiencies in adrenal steroidogenesis, most commonly due to 21-hydroxylase deficiency (21OHD), which accounts for over 95% of cases (1). The condition results in impaired cortisol synthesis and, in the classic forms, often affects aldosterone production as well. As a compensatory response, elevated adrenocorticotropic hormone (ACTH) levels stimulate adrenal androgen excess, leading to variable degrees of virilization and metabolic disturbances depending on the severity of the enzymatic defect (2).

The clinical spectrum of CAH due to 21OHD ranges from the severe salt-wasting (SW) and simple virilizing (SV) forms, typically presenting in infancy, to the milder nonclassical (NC) phenotype, which may manifest later in childhood or adolescence with signs of androgen excess. The incidence of classic CAH is estimated at 1:14,000 to 1:18,000 live births worldwide, although frequencies vary by population and genetic background (3). Advances in neonatal screening and standardized treatment regimens have improved outcomes, but patients continue to face significant challenges related to growth, fertility, and long-term metabolic health.

The clinical phenotype of congenital adrenal hyperplasia (CAH) due to 21-hydroxylase deficiency is primarily determined by the degree of residual enzymatic activity encoded by the CYP21A2 gene variants. Multiple large-scale studies have established a robust genotype–phenotype correlation, especially for severe mutations. Riedl et al. demonstrated that genotypes classified as null or Group A (with 0–1% residual activity) predict the salt-wasting phenotype with high accuracy (97% and 91%, respectively). In comparison, genotypes B and C exhibit greater phenotypic variability and reduced concordance (46% and 58%) (4). Similarly, New et al. confirmed high genotype–phenotype concordance rates in a cohort of over 1,500 families, noting that the less severe alleles (especially V281L) were often associated with unexpected classical presentations (5). The predictive validity of genotype classification also depends on clinical context, as illustrated by studies showing that phenotypes assigned to newborns through screening programs tend to be more severe than expected for their genotype, possibly reflecting clinical caution in early treatment (6).

Despite extensive evidence supporting genotype–phenotype correlations in CAH, most published data originate from European or North American cohorts, and limited information is available from Latin American populations. Few studies have examined the potential influence of mutational burden, the total number of pathogenic variants identified, on the accuracy of phenotype prediction. In this study, we aimed to characterize the clinical and molecular profile of patients with CAH followed in a specialized rare disease program in Colombia. We also evaluated the concordance between predicted genotype-based enzymatic activity and clinical phenotype, and explored whether mutational burden and sex modify the accuracy of genotype–phenotype prediction.

## Methods

We conducted a cross-sectional study using anonymized electronic medical records from patients enrolled in the Rare Diseases Program at Hospital Pablo Tobón Uribe (MedellÍn, Colombia). This institutional program provides specialized care to approximately 3,600 individuals with rare and ultra-rare conditions. For this analysis, we included all patients with a confirmed diagnosis of congenital adrenal hyperplasia (CAH). The dataset included demographic characteristics, biochemical profiles, imaging results, treatment modalities, and clinical follow-up data. Data was collected between March/01 and July/31 2024.

Descriptive analyses were performed using absolute and relative frequencies for categorical variables, and measures of central tendency and dispersion for continuous variables. Comparisons between groups were conducted using the chi-square or Fisher’s exact test for categorical variables, and the Kruskal–Wallis or Wilcoxon rank-sum test for continuous variables, as appropriate.

Patients with available genetic data were stratified according to a four-tier genotypic classification system based on predicted residual 21-hydroxylase enzymatic activity, consistent with established genotype–phenotype correlation frameworks. The categories included: Null (no residual activity), Group A (<1% activity), Group B (2–11%), and Group C (20–50%).

The relationship between genotype-predicted enzymatic activity and clinical phenotype was assessed by comparing the predicted genotype category with the observed clinical presentation, categorized into three groups: salt-wasting (SW), simple virilizing (SV), and nonclassical (NC). Concordance was defined as an exact match between genotype prediction and clinical phenotype.

Mutational burden was quantified as the total number of pathogenic variants identified per individual. For the primary analysis, it was dichotomized as “low” (≤2 mutations) and “high” (>2 mutations). A binary logistic regression model was fitted with concordance as the dependent variable and mutational burden as the predictor. Given the presence of sparse cells and quasi-complete separation, Firth’s penalized logistic regression was applied to improve model stability and reduce bias. A secondary model including an interaction term between sex and mutational burden was also fitted. Odds ratios (ORs) and 95% confidence intervals (CIs) were calculated.

All analyses were performed in R version 4.3.1. Statistical significance was defined as *p* < 0.05. This study was approved by the Institutional Ethics Committee of Hospital Pablo Tobón Uribe (approval code: 10/2023) and complied with national and international ethical standards. All data were anonymized to preserve patient confidentiality.

## Results

A total of 75 patients with confirmed adrenal hyperplasia were included. The median age was 19.2 years (SD: 11.5), with a female predominance (66.7%). Baseline clinical and biochemical characteristics are summarized in Table 1. Regarding treatment history, 98.7% had received dexamethasone, 76.0% prednisolone, 41.3% fludrocortisone, and 29.3% hydrocortisone at some point.

Among pediatric patients (n = 38), the mean height-for-age Z-score was 3.13 (SD: 3.82). When stratified by phenotype, Z-scores were 2.52 (SD: 4.30) in SW, 2.56 (SD: 3.29) in SV, and 4.82 (SD: 2.38) in NC forms. Although NC patients showed higher mean scores, differences were not statistically significant (*p* = 0.271). In adults, the mean normalized height Z-score was –0.90 (SD: 1.07), with significantly lower values in males (–1.72, SD: 0.60) compared to females (–0.63, SD: 1.06) (*p* = 0.0061).

The mean BMI among children was 20.1 kg/m^2^ (SD: 4.5), with a mean BMI-for-age Z-score of 3.80 (SD: 2.86), indicating a higher-than-expected distribution. In adults, the mean BMI was 26.3 kg/m^2^ (SD: 4.9), consistent with the overweight range.

Genetic testing was available for 48 patients. The most frequent pathogenic variant was p.Val282Leu (56.2%), followed by p.Leu308Phe and c.293_13cg (29.3%). Less common mutations included p.Ile173Asn (17.3%), c.293_13ag (12.0%), and p.Arg357Trp (5.3%). Large deletions or gene conversions were not detected. Most patients had multiple pathogenic variants: 60.4% had two, 35.4% had three, and 4.2% had four or more (Table 2). The distribution and relative frequency of CYP21A2 mutations among genotyped individuals are depicted in Figure 2. Mutations are ordered by genomic position, and their color reflects the predicted residual enzymatic activity.

**Figure 1.**
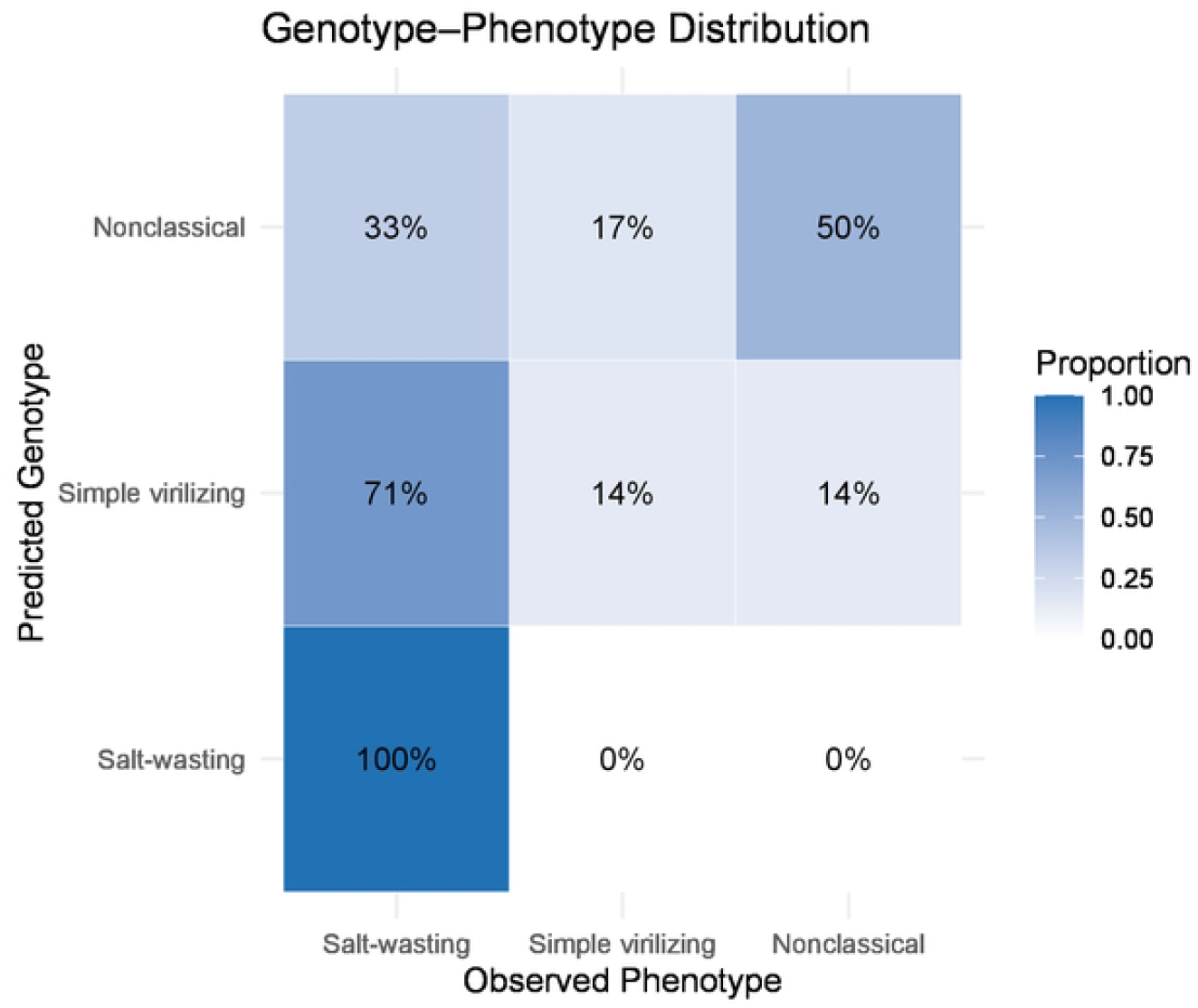
Distribution of Observed Phenotype by Predicted Genotype in Patients with Congenital Adrenal Hyperplasia. The heatmap illustrates the proportional distribution of clinical phenotypes (columns) across predicted genotype classes (rows) in 48 individuals with congenital adrenal hyperplasia due to 21-hydroxylase deficiency. Each cell represents the proportion of patients with a given predicted genotype (Salt-wasting, Simple virilizing, or Nonclassical) who were clinically classified with a corresponding phenotype. Color intensity reflects proportion, with darker blue indicating higher concordance. Predicted genotypes were derived from pathogenic variant interpretation, following standard genotype-to-phenotype prediction models.

**Figure 2.**
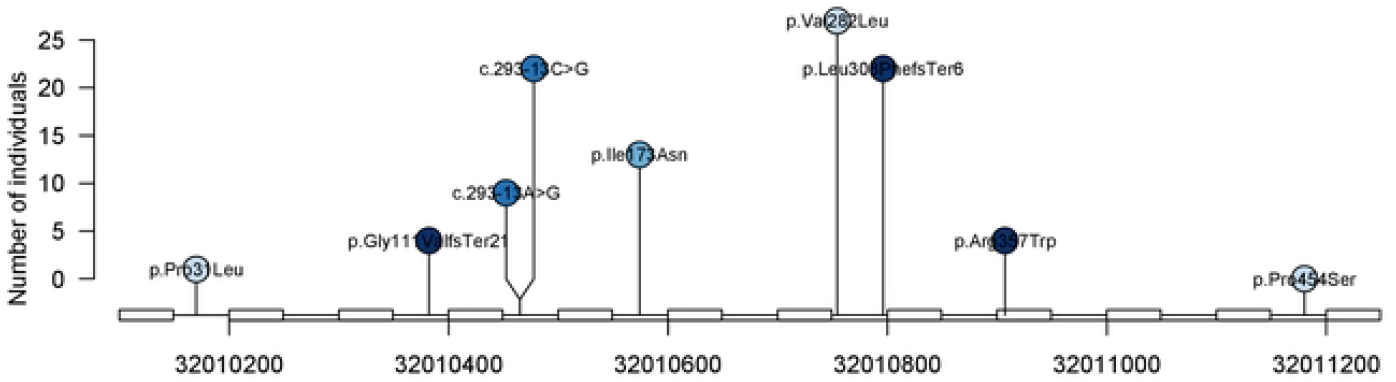
Distribution of CYP21A2 pathogenic variants Identified in patients with congenital adrenal hyperplasia. Lollipop plot representing the position and frequency of mutations across the CYP21A2 gene (chromosome 6). Circle size is proportional to the number of individuals harboring each variant. Colors indicate predicted residual 21-hydroxylase enzymatic activity: dark blue (0%), medium blue (<1%), light blue (2−11%), and pale blue (20−50%). The most frequent variant was p.Val282Leu (n = 27), followed by p.Leu308PhefsTer6 and c.293-13C>G (n = 22 each). One patient may harbor more than one mutation. Only mutations with known genomic coordinates and predicted functional classification are shown.

Based on the four-group genotypic classification, 56.2% were assigned to Group C, 29.2% to Group A, 12.5% to Group B, and 2.1% to Null. Genotype–phenotype concordance was higher among more severe genotypes: all Null and 85.7% of Group A individuals presented with the SW phenotype. Conversely, individuals in Groups B and C showed greater phenotypic variability. Notably, 66.7% of Group B and 55.5% of Group C patients exhibited phenotypes more severe than predicted. Figure 1 displays the distribution of clinical phenotypes across genotype categories.

Among individuals with SW-predicted genotypes, 63% displayed a concordant clinical phenotype, while 27% were classified as NC. In contrast, among NC-predicted genotypes, only 55% exhibited the expected phenotype.

In the penalized logistic regression model, individuals with high mutational burden had significantly lower odds of genotype–phenotype concordance (OR = 0.18, 95% CI: 0.03–0.94, *p* = 0.041). In the interaction model, neither sex nor mutational burden, nor their interaction, was significantly associated with concordance (interaction term: OR = 0.25, 95% CI: 0.001–61.7, *p* = 0.55). While results suggest a trend toward reduced predictive accuracy with higher mutational burden, no effect modification by sex was detected.

## Discussion

In this cross-sectional study of patients with congenital adrenal hyperplasia (CAH) followed at a specialized rare disease center in Colombia, we confirmed genotype–phenotype correlations consistent with those reported in previously published cohorts. As observed in larger European and North American studies, genotypes associated with minimal or absent residual enzymatic activity (Null and Group A) were strongly linked to the salt-wasting (SW) phenotype, while milder genotypes (Groups B and C) demonstrated greater clinical variability. Our findings reinforce the validity of the four-tier genotypic classification framework for predicting disease severity, while also highlighting its limitations, particularly in the context of milder alleles and increased mutational burden. Notably, more than one-third of patients in Group C exhibited clinical presentations more severe than expected.

In contrast to our results, the CaHASE study (6) reported limited genotype–phenotype concordance in a UK adult cohort with CAH. Using a similar classification system based on predicted residual enzymatic activity, the study observed substantial overlap in clinical presentations across genotypic groups, suggesting that genotype alone was a poor predictor of adult phenotype. In our cohort, however, concordance remained relatively high for severe genotypes (Null and Group A) and declined progressively with increasing residual enzymatic activity, consistent with previously published pediatric data.

Our findings of partial genotype–phenotype concordance, particularly among individuals with Group B and C genotypes, are consistent with the international cohort reported by New et al., which analyzed over 1,500 families with CAH due to 21-hydroxylase deficiency. In that study, concordance was observed in fewer than 50% of the 45 genotypes evaluated, with substantial variability, especially among individuals carrying the I172N and V281L mutations. Mutations typically associated with the simple virilizing phenotype, such as I172N, were frequently linked to salt-wasting presentations, while V281L—often linked to nonclassical CAH—was also found in individuals with classic disease. These findings align with our observation that over half of patients in Group C exhibited phenotypes more severe than predicted, reinforcing the notion that additional genetic or epigenetic modifiers may influence clinical presentation beyond the primary CYP21A2 mutations.

In our Colombian cohort, genotype–phenotype concordance followed the expected gradient, with high agreement among severe genotypes (Null: 100%; Group A: 85.7%) and diminishing concordance in milder categories (Group B: 33.3%; Group C: 44.4%). This trend mirrors the findings of Riedl et al. (4), who reported genotype–phenotype concordance rates of 97% and 91% for Null and Group A genotypes, respectively, and lower concordance for Groups B (46%) and C (58%). As in our study, their results highlighted that individuals harboring milder mutations frequently presented with more severe clinical phenotypes, underscoring the limitations of relying solely on genotypic classification in individuals with intermediate residual enzymatic activity. These observations support the need for integrating additional clinical and biochemical parameters when evaluating disease severity and counseling patients.

Structure-based modeling has emerged as a valuable strategy to enhance the predictive power of genotype–phenotype correlations in 21-hydroxylase deficiency. Bruque et al (7). demonstrated that computational estimates of residual enzymatic activity based on protein stability predictions (ΔΔG) show strong agreement with clinical phenotype and in vitro functional assays. This method offers a promising complement to traditional genotypic classification, particularly for rare or uncharacterized variants and compound heterozygotes with uncertain phenotypic outcomes. Our finding of reduced genotype–phenotype concordance among individuals with higher mutational burden may reflect the limitations of sequence-based prediction alone. Integrating structure-informed models could improve phenotype prediction in clinical settings where functional assays are unavailable or phenotypic variability is pronounced.

In classical genotype–phenotype models of CAH, the less severe of the two CYP21A2 mutations is typically assumed to determine the predicted clinical phenotype, based on the idea that the milder allele largely governs residual enzymatic activity. However, our findings challenge this paradigm. We observed that individuals harboring more than two pathogenic variants had significantly lower odds of genotype–phenotype concordance. This suggests that increased mutational burden may introduce greater complexity in phenotype expression, possibly due to additive effects, cis–trans interactions, or unrecognized genetic modifiers. The observed reduction in predictive accuracy emphasizes the limitations of traditional biallelic models in complex genotypes. It supports the incorporation of total mutational load as an additional parameter in clinical interpretation and genetic counseling.

This study has several limitations. First, the sample size, particularly in some genotype subgroups, was relatively small, limiting the statistical power to detect subtle associations and increasing the risk of type II error. Second, the classification of clinical phenotype was based on retrospective chart review, which may be subject to misclassification bias, particularly in cases with incomplete documentation or evolving phenotype over time. Third, the analysis did not include functional validation of identified variants, and genotype–phenotype associations were inferred solely from predicted enzymatic activity, which may not capture the full spectrum of pathogenicity. Fourth, although we considered mutational burden, we did not explore the potential impact of variant phase (cis vs. trans), structural rearrangements, or deep intronic mutations that could influence gene expression. Finally, our cohort is derived from a single center in Colombia and may not be generalizable to broader populations; nevertheless, it provides crucial insights into CAH in underrepresented Latin American populations.

This study also has important strengths. It represents one of the few genotype–phenotype correlation studies conducted in a Latin American population, contributing valuable data to a region historically underrepresented in CAH research. The cohort was derived from a well-defined institutional program for rare diseases, ensuring standardized clinical follow-up and access to detailed medical records—the integration of clinical, biochemical, and genetic data allowed for a comprehensive characterization of patients. Furthermore, the application of advanced statistical methods, including Firth’s penalized logistic regression, addressed potential bias due to small sample size and sparse data, enhancing the robustness of the genotype–phenotype concordance analysis. Finally, the study introduced the concept of mutational burden as a novel and potentially informative factor influencing the accuracy of genotype-based phenotype prediction, which may inform future diagnostic and counseling strategies.

## Conclusions

In this cohort of Colombian patients with congenital adrenal hyperplasia, we confirmed the relevance of genotype–phenotype correlations based on predicted enzymatic activity. Severe genotypes (Null and Group A) showed high concordance with the salt-wasting phenotype, while milder genotypes (Groups B and C) exhibited greater clinical variability. Notably, an increased number of pathogenic variants was associated with reduced concordance, suggesting that mutational burden may affect phenotype expression beyond the influence of the milder allele alone. These findings support the utility of genotype-based classification while underscoring the need to consider total mutational load and integrate complementary clinical and molecular data in phenotype prediction.

## Data Availability

The dataset analyzed in this study is held by the institution that approved the research protocol. Data may be made available upon reasonable request and with prior authorization from the Institutional Ethics Committee.

## Ethical implications

This study was approved by the Institutional Ethics Committee of Hospital Pablo Tobón Uribe (approval code: 10/2023). All data were obtained from existing electronic medical records and analyzed using a fully anonymized database. No direct patient contact occurred, and no identifiable personal information was collected or stored. Privacy and confidentiality were strictly maintained throughout the study in accordance with national and international ethical standards for research involving human data.

## Acknowledgements

None

## Authors’ contributions

Carlos E. Builes-Montaño, Gustavo A. Giraldo Ospina, and Carolina Baquero Montoya contributed to the conceptualization of the study. Ana M. Perilla-Espinal, Valentina Zapata-López, and Sofía Villada-Montoya were responsible for data collection. Carlos E. Builes-Montaño performed the statistical analysis. All authors participated in the drafting and critical revision of the manuscript, contributed to the study design and interpretation of results, and approved the final version for submission.

## Use of Artificial Intelligence

Artificial intelligence tools were employed to support specific phases of the manuscript development. Grammarly assisted with grammar and style revision during the final editing phase. No generative AI tools were used to produce original scientific content, interpret results, or replace the authors’ critical analysis and writing responsibilities.

